# Post-Viral Frailty in Long COVID: A Distinct Phenotype within Veterans

**DOI:** 10.1101/2025.02.04.25321653

**Authors:** Jerry Bradley, Elizabeth Bast, Natasha M. Resendes, Fei Tang, Victor D. Cevallos, Dominique M. Tosi, Leonardo Tamariz, Ana Palacio, Iriana S. Hammel

**Affiliations:** Department of Medicine, University of Miami Miller School of Medicine, Miami, Florida, USA; Miami Veterans Administration (VA) Healthcare System, Department of Ambulatory Medicine, Miami FL; Miami Veterans Administration (VA) Healthcare System Geriatric Research, Education, and Clinical Center (GRECC), Miami, Florida, USA; Miami Veterans Administration (VA) Healthcare System, Department of Medicine, Miami FL

## Abstract

**Background:** Long COVID is characterized by persistent symptoms affecting one or more organ systems for at least 3 months following a SARS-CoV-2 infection. The pathophysiologic mechanisms of this complex disease are poorly understood. Beyond the described symptoms of fatigue, dyspnea, myalgias, among others, Long COVID can affect the patient’s ability to work and function in society compared to their baseline. Frailty is defined as the decline of physiologic reserve that leads to increased vulnerability to stressors and poor health outcomes. Our study aimed to examine the characteristics of frailty seen in patients with Long COVID compared to the frailty seen in aging patients with multimorbidity.

**Methods:** This is a retrospective cohort study conducted in the Miami Veterans Affairs Medical Center (VAMC). The data used to calculate the Fried phenotype through the Johns Hopkins frailty calculator was collected from two separate clinics, a Long COVID clinic and a Geriatric Frailty clinic. We obtained the VA Frailty Index from VA CDW (Corporate Data Warehouse).

**Results:** We included 106 patients from the Long COVID clinic and 97 from the frailty clinic. Patients from the Long COVID clinic were significantly younger than those from the frailty clinic (60±12.6 vs.. 79.8±5.8, p<0.01). In the standard frailty group, weakness and slowness were the predominant features present in both the frail and pre-frail groups, with increasing exhaustion and lower activity in the frail group. Patients with frailty in the Long COVID group experienced exhaustion and low activity at a higher rate than those in the Geriatric frailty clinic.

**Conclusions:** Long COVID may predispose patients to develop a subtype of frailty (“post-viral frailty”) that presents with a higher frequency of exhaustion and low activity. This frailty appears phenotypically different from the frailty encountered in geriatric patients with multimorbidity, which presents more often with slowness and weakness as the initial drivers.

## Introduction

Long COVID is a complex disease process with poorly understood pathophysiological mechanisms[1]. Per the 2024 NASEM Long COVID Definition, Long COVID (LC) is an infection-associated chronic condition (IACC) that occurs after SARS-CoV-2 infection and is present for at least 3 months as a continuous, relapsing and remitting, or progressive disease state that affects one or more organ systems [2]. Although symptoms are wide-ranging, a meta-analysis found that fatigue, shortness of breath, olfactory dysfunction, and myalgia are among the most common symptoms reported[3]. Beyond these manifestations, in some cases, Long COVID fatigue can cause reduced physical and mental activity, limiting a patient’s ability to work and function in society compared to their pre-infectious baseline[4]. Unfortunately, symptoms may last for years, with active research currently investigating the long-term sequelae of this disease process[1].

Frailty has traditionally been defined as a decline in the physiological reserve of patients, leading to a vulnerability to external stressors linked to poor outcomes[5]. Similar to Long COVID, frailty can impact function and work productivity [6]. Two major models are commonly used to determine frailty in patients. The first is the Fried phenotype, based on five physiological factors: weakness measured by grip strength, slowness measured by gait speed, weight loss, and self-reported low physical activity and exhaustion.[7]. These characteristics can help providers identify appropriate interventions while monitoring the response to therapy in repeat assessments. The second model is based on deficit accumulation as a predictor of frailty and its outcomes[8]. Under this model, the accumulation of health deficits, such as chronic medical conditions, increases the risk of frailty and death[9]. One of the advantages of using the deficit accumulation model, commonly represented as a frailty index, is that it does not require a physical assessment of the patient and has been well-validated to predict mortality in multiple studies[10]. Rather than seeing these as competing frailty models, experts have suggested that both are complementary to understanding the complexity of frailty using different approaches, with each yielding valuable clinical insights[11].

Although the Frailty phenotype and Frailty indices have only been validated in patients over 65 years old [7, 12] there have been multiple attempts to use these calculators with younger patients, and we recognize that younger people may also develop frailty. A large review identified many papers that included populations of people over and under 60 years that demonstrated predictive validity for mortality and/or hospital admissions of both the frailty phenotype and frailty indices [13]. Our study aimed to examine the development of frailty in patients with Long COVID compared to the frailty seen in aging patients with multimorbidity. Our goal was to evaluate frailty in each of these cohorts using both the Fried phenotype through the validated Johns Hopkins frailty calculator and the deficit accumulation model through the Veterans Affairs Frailty Index (VA-FI)[14]. We propose that the post-viral frailty that occurs in Long COVID patients is a unique phenotypic presentation, with low physical activity and exhaustion being the predominant characteristics. We also propose that the deficit accumulation model may not be as well-suited for diagnosing frailty in Long COVID due to the post-viral pathophysiology causing the emergence of a frailty state rather than the accumulation of multiple health conditions through the aging process.

## Methods

### Study Design

The frailty clinic cohort was comprised of patients seen in the Geriatric Research, Education, and Clinical Center (GRECC) Frailty Clinic. The GRECC Frailty Clinic evaluates patients over 65 years old who are having difficulty with daily activities, mobility, and/or cognition, among other problems. Clinic enrollment occurs by referral from their PCP or other providers. Patients in the clinic are evaluated using the frailty phenotype by completing the Johns Hopkins (JH) frailty assessment calculator.

The Long COVID cohort comprised patients seen in the Miami VA Long COVID clinic. The Miami VAMC Long COVID clinic evaluates patients struggling with new or worsened symptoms lasting at least 12 weeks after a COVID-19 infection. Clinic enrollment occurs by referral and through a digital screening program. Starting in January 2024, as part of a clinical innovation project, all patients seen in person at the Miami VAMC clinic were screened for frailty by completing the JH frailty phenotype assessment as described above. The Miami Veterans Affairs Healthcare System Institutional Review Board approved this study. The IRB granted a waiver for informed consent. The data was fully anonymized according to VHA standards prior to analysis. The data was accessed on 01/09/2024 to 17/01/2025.

### Data Source

We calculated VA Frailty Index (VA-FI) for each patient using data from VA CDW (Corporate Data Warehouse) at the time they presented to the respective clinic. The 31-item VA Frailty Index (VA-FI)^15^ was developed based on the deficit accumulation conceptual framework (Table s1). We categorized patients based on the score as Severe Frail (0.40 or above, Frail <u>(></u>0.21), Pre-frail (0.11-0.20), or robust (<0.10). Demographic information was collected via review from the VA EHR and from COVID-19 Shared Data Resource [15].

### Statistical analysis

Continuous variables were presented as mean ± standard deviation; categorical variables were presented as frequency and percent. We compared numerical variables using t-tests and compared categorical variables using Chi-squared test. All the statistical analyses were performed with R (the R project for statistical computing, version 4.0.5).

## Results

We included 106 patients from the Long COVID clinic and 97 from the frailty clinic and presented the patients demographic in Table 1. Patients from the Long COVID clinic were significantly younger than those from the frailty clinic (60±12.6 vs.. 79.8±5.8, p<0.01). There are more Hispanic patients in the Long COVID clinic compared to the frailty clinic,28 (26.9%),vs.. 13 (13.4%).

**Table 1:** Cohort demographic

|  | **Long COVID**<br>**N=106** | **Frailty Clinic**<br>**N=97** | **p-values** |
| --- | --- | --- | --- |
| Male Gender | 82 (78.9%) | 94 (96.9%) | <0.01 |
| Age | 60±12.6 | 79.8±5.8 | <0.01 |
| Age Groups, n (%) |  |  |  |
| <55 | 31 (29.8%) | 0 |  |
| 55-64 | 33 (31.7%) | 0 |  |
| 65-74 | 25 (24.0%) | 13 (13.4%) | <0.01 |
| 75-84 | 13 (12.5%) | 63 (63%) |  |
| ≥85 | 2 (1.9%) | 21 (21%) |  |
| Race, n (%) |  |  |  |
| White | 51 (49%) | 56 (57.7%) |  |
| Black | 43 (41.3%) | 37 (38.1%) | 0.71 |
| Asian | 2 (1.9%) | 0 |  |
| American Indian or<br>Alaska Natives | 0 | 0 |  |
| Native Hawaiian or<br>Other Pacific<br>Islander | 3 (2.9%) | 0 |  |
| Unknown | 4 (3.8%) | 4 (4.1%) |  |
| Ethnicity, n (%) |  |  |  |
| Hispanic | 28 (26.9%) | 13 (13.4%) |  |
| Not Hispanic | 74 (71.2%) | 81 (83.5%) | 0.04 |
| Unknown | 1 (1.0%) | 3 (3.1%) |  |

There was a significant difference in the identification of frailty between groups using the VAFI model but not with the Fried model. These two models demonstrated different assessments of frailty as 66% of the LC model was considered frail by VAFI, while only 26% by Fried, and among the patients in the frailty clinic, 93% were frail per VAFI and only 39% by Fried (Table 2).

**Table 2:**
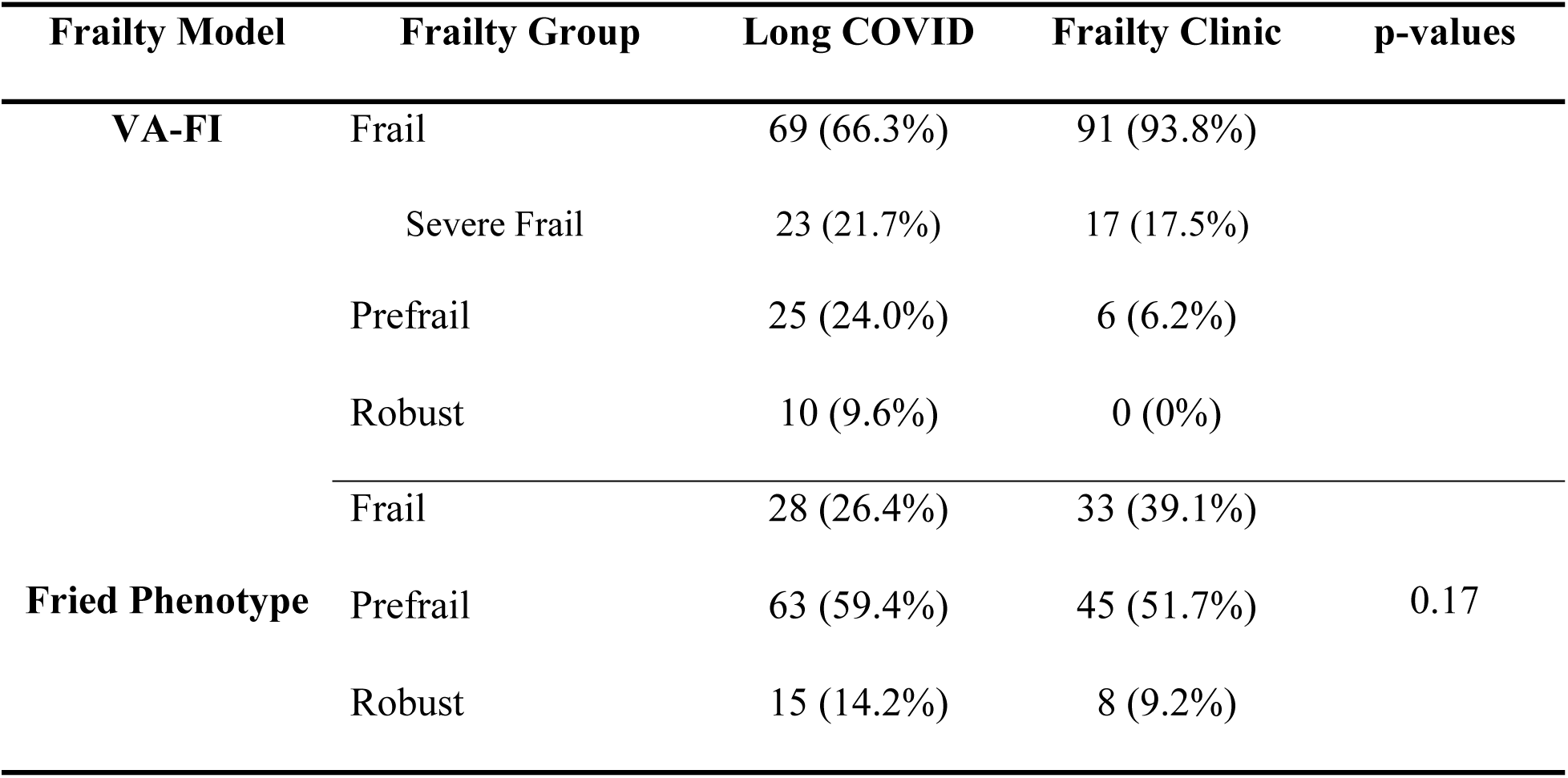
Frailty percentage by group and cohort type using the Veteran Affairs Frailty Index (VA-FI) and Fried Phenotype

Among frail patients (Table 3), those in the Long COVID group had higher rates of exhaustion (96.4%) and low activity (78.6%) compared to the Frailty clinic group (exhaustion 57.6%, low Activity 63.6%). Conversely, Frailty clinic patients had higher rates of weakness (97%) and slowness (87.9%) compared to the Long COVID group (weakness 75% and slowness 46.4%). However, the overall differences between these group components approached but did not reach significance (p = 0.08). Among the prefrail patients, a similar pattern was seen with Long COVID patients having higher rates of exhaustion (68.3% vs. 12.8%) and low activity (28.6 vs. 10.3%) as compared to the frailty clinic. In addition, frailty clinic patients had higher rates of weakness (56.4% vs 27%) and slowness (51.3% vs 9.5%). Within the prefrail group, there was a significant difference between the Long COVID and frailty clinic phenotype components (p < 0.01).

**Table 3:** Rate of presentation by Fried phenotype category in frail and pre-frail patients from both cohorts.

|  | Long COVID Clinic | Frailty Clinic | P-values |
| --- | --- | --- | --- |
|  | Frail<br>(N=28) | Frail<br>(N=33) |  |
| **Weakness** | 21 (75%) | 32 (97.0%) |  |
| **Slowness** | 13 (46.4%) | 29 (87.9%) | 0.08 |
| **Weight loss** | 9 (32.1%) | 9 (27.3%) |  |
| **Exhaustion** | 27 (96.4%) | 19 (57.6%) |  |
| **Low Activity** | 22 (78.6%) | 21 (63.6%) |  |
|  | Prefrail<br>(n=63) | Prefrail<br>(n=39) |  |
| **Weakness** | 17 (27.0%) | 22(56.4%) |  |
| **Slowness** | 6 (9.5%) | 20(51.3%) |  |
| **Weight loss** | 3 (4.8%) | 10(25.6%) | <0.01 |
| **Exhaustion** | 43(68.3%) | 5(12.8%) |  |
| **Low Activity** | 18(28.6%) | 4(10.3%) |  |

## Discussion

### Differences in Phenotype Between Groups

In the patients evaluated in the GRECC frailty clinic (standard frailty groups), weakness and slowness were the predominant features in both the frail and pre-frail groups, with increasing exhaustion and lower activity in the frail group. Frailty in older adults is thought to be driven by weakness and reduced muscle strength, with sarcopenia often developing as a precursor to frailty [16]. Studies have shown that decreased muscle strength is highly predictive of frailty status over several years [17].

Patients with frailty in the Long COVID group experienced exhaustion and low activity at a higher rate than those in the Geriatric frailty clinic. This relationship was even more pronounced in the pre-frail category. These findings were anticipated, given the pathophysiological drivers of Long COVID within each frailty category. Fatigue is a dominant complaint of individuals with Long COVID, which can last for months to years at a time [18]. Additionally, studies have shown evidence of lower physical activity and impaired function, which significantly affect the quality of life of patients with Long COVID [19]. As a result, patients with post-viral conditions, such as Long COVID, have a higher probability of exhaustion and lower physical activity phenotype, as seen in our results. These differences in phenotype presentation may be partly driven by the underlying pathophysiological mechanisms, response to exertion, and treatment outcomes.

Several of the patients from the Long COVID clinic are younger than 65 and we may in fact be identifying the most frail patients in this group, as they are meeting criteria for frailty in older patients – meaning they are worse than would be expected of an older adult – despite their younger age. Some patients who don’t meet the criteria may have a falsely “normal” gait speed and grip strength that could be abnormal if we were looking at age-based norms.

### Post Viral Frailty as a Distinct Phenotype Presentation

In our study, we propose the term post-viral frailty to refer to patients who develop pre-frailty or frailty after a viral illness. Long COVID represents a unique post-viral condition. As discussed previously, both frailty and Long COVID share disruptions in metabolic, inflammatory, endocrinologic, and immunologic pathways. However, the effects of these disruptions, their primary drivers, and their long-term impacts on health and function as they relate to frailty remain an active area of investigation. We propose post-viral frailty as a distinct phenotype of frailty that requires further investigation to define its pathophysiological cause, understand its progression, and identify appropriate treatment strategies.

From a pathophysiological perspective, frailty is thought to develop from disruption of the immune and inflammatory systems involving skeletal muscle function, endocrinological issues, and cellular changes resulting in oxidative stress [20]. Frailty, driven by these disruptions affecting gait and strength, has physical therapy as one of the mainstays of treatment [21]. A comprehensive meta-analysis found that structured intervention programs targeting muscle strengthening are beneficial for frail older adults [22]. In addition to improving physical function, exercise is also believed to be productive in reducing some drivers of frailty, such as oxidative damage and chronic inflammation, while improving mitochondrial function and endocrine pathways [23].

Long COVID shares many of these pathophysiological characteristics, with mitochondrial dysfunction[24], chronic inflammation[25], and alterations in immune functions being some of the predominant features [26]. Long COVID is believed to share many similarities with myalgic encephalomyelitis/chronic fatigue syndrome (ME/CFS)[27]. Patients with both conditions can experience post-exertional malaise (PEM) [28]. Post-exertional malaise occurs after an individual exerts themselves beyond a threshold level, causing increased fatigue and exhaustion. A recent study showed that patients with post-exertional malaise in Long COVID had structural skeletal muscle changes and local metabolic disturbances [29]. This effect is paradoxical in that physical activity increases and worsens the disease process rather than being beneficial. Numerous studies before COVID-19 observed in ME/CFS cohorts warned against physical activity as it could be harmful [30, 31]. Therefore, post-viral frailty, as developed in Long COVID patients who have PEM causing low physical activity and exhaustion, may not benefit from standard treatment approaches to frailty. Instead, patients, as seen in our cohort, were treated with modified physical therapy that involved pacing, diaphragmatic breathing and autonomic rehabilitation. Current evidence shows that pacing can improve outcomes in Long COVID [32] by helping patients understand the limits of their physical activity and prevent PEM. Although Long COVID patients share phenotypic characteristics with geriatric patients with frailty, further research is required to evaluate the optimal treatment options for post-viral frailty beyond traditional interventions.

### Limitations of the Deficit Accumulation Model in Post-Viral Frailty

The deficit accumulation model of frailty was developed based on the theory that, as a function of aging, multiple health deficits contribute to the risk of developing frailty. Individuals with significant health deficits or an accumulation of them are at a higher risk than other patients of equivalent age [33]. Based on the frailty index that was developed, patients can be stratified according to their frailty status based on accumulated deficits [12]. The Veterans Affairs Frailty Index (VA-FI), created on this model, is a well-validated clinical tool for Veterans aged over 65 [34]. In the cohort of patients referred to the frailty clinic, 100% were either frail or pre-frail, vastly outstripping the prevalence in the LC group; however, in the setting of post-viral frailty, the underlying pathophysiological assumption behind the deficit accumulation model may not be as applicable. Individuals with post-viral frailty in the Long COVID group acquired a viral illness, which resulted in changes in immunological, metabolic, and other systemic responses [35]. Their frailty presentation was not brought about by an accumulation of deficits through the aging process, but rather as a response to an infectious process. Although the precise pathophysiological mechanisms underlying Long COVID remain elusive [36], individuals in the Long COVID cohort were healthier and younger than their counterparts in the frailty comparison group. As seen between the Long COVID and frailty clinic groups, there was a wide degree of variation between the frailty group stratifications, reflected in the fewer chronic medical conditions present in the Long COVID cohort. Therefore, the underlying clinical model regarding the accumulation of health conditions as a marker for frailty, although validated and clinically useful in older adults with frailty, may not be applicable to patients with post-viral frailty because of the post-infectious sequelae that give rise to the post-viral frailty state in previously robust patients.

### Strengths and Limitations

One of the major strengths of this study is the use of both the deficit accumulation model and the Fried phenotype to understand the frailty characteristics within both groups. Currently, there are limited data regarding this physical frailty phenotype in Long COVID patients, as many studies have been conducted using only the deficit accumulation model. Other strengths of this study include the use of a comparative frailty cohort. Given the design of the study, we were able to analyze frailty characteristics across multiple patient groups within the Veteran community.

Limitations of the study include the lack of validation of frailty phenotypic measures in younger populations and the lack of a non-Long COVID control group with similar age and comorbidity distribution for comparison of frailty in the general population using these methods. Another limitation is potential for self-selection: clinic patients were referred by their primary care providers either to the frailty clinic or the Long COVID clinic within the VA health system. Patients who utilized this referral may have had more disabling symptoms or concerns, requiring further specialty evaluation. As a result of this self-selection process, the cohort of patients present within this study may not accurately represent the more general populations of patients with Long COVID or frailty. Another limitation was the lack of longitudinal assessments. Data on frailty characteristics were collected at a single point during the study. Although this is beneficial for understanding the phenotypic presentation of frailty within both groups, further research is required to understand the changes in frailty characteristics over time as a function of specific interventions.

## Conclusion

Post-viral frailty in patients with Long COVID represents a distinct phenotype, with exhaustion and low physical activity being the predominant drivers. Further research is required to understand intervention strategies that may be beneficial for these patients.

## Data Availability

The datasets presented in this article are not readily available because of the Department of Veterans Affairs data policies. Requests to access the datasets should be directed to the Department of Veterans Affairs

## Supplemental Table

**Table S1:**
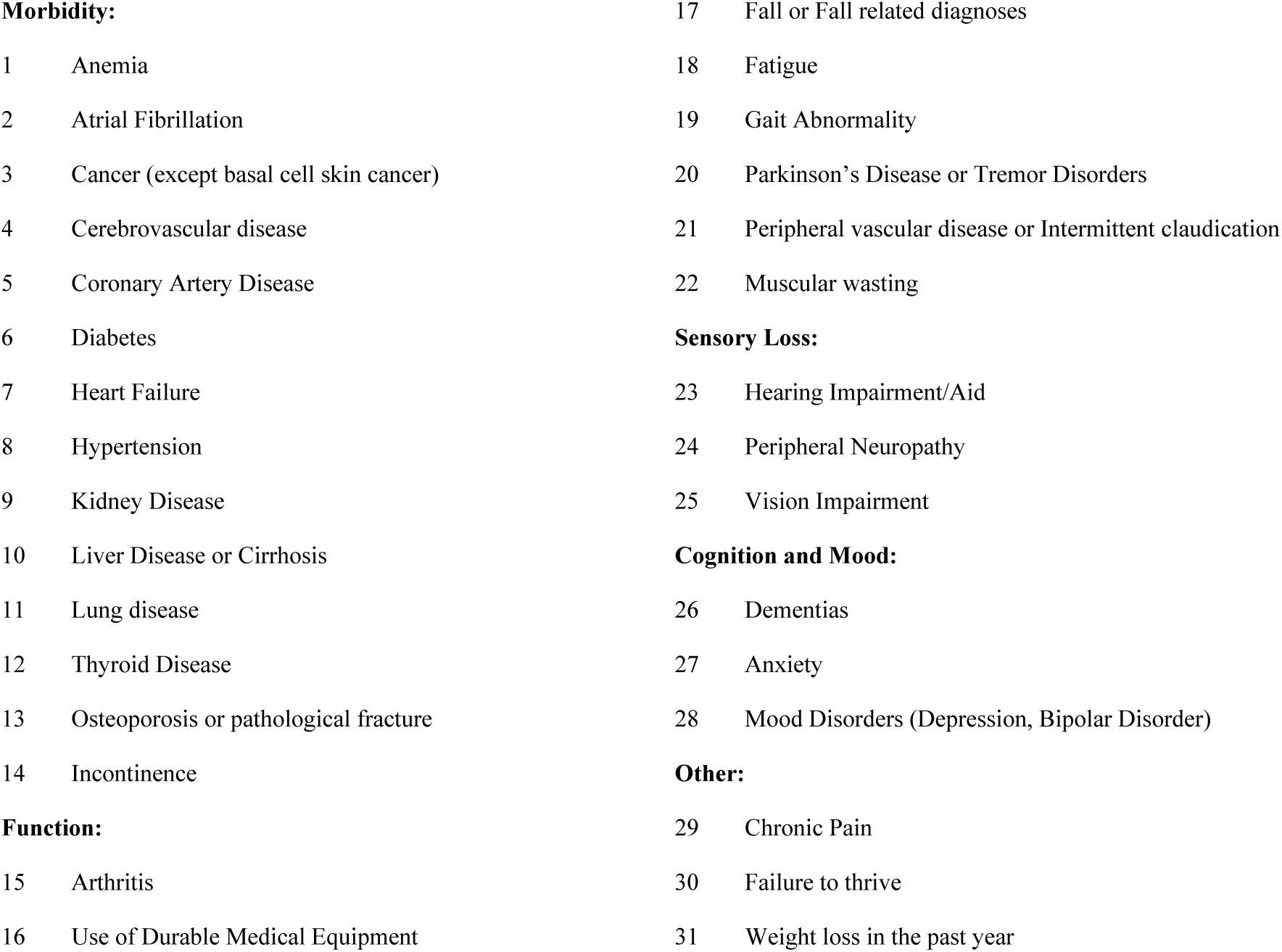
VA Frailty Index Variables

|  |  |  |  |
| --- | --- | --- | --- |
| **Morbidity:** |  | 17 | Fall or Fall related diagnoses |
| 1 | Anemia | 18 | Fatigue |
| 2 | Atrial Fibrillation | 19 | Gait Abnormality |
| 3 | Cancer (except basal cell skin cancer) | 20 | Parkinson’s Disease or Tremor Disorders |
| 4 | Cerebrovascular disease | 21 | Peripheral vascular disease or Intermittent claudication |
| 5 | Coronary Artery Disease | 22 | Muscular wasting |
| 6 | Diabetes | **Sensory Loss:** |  |
| 7 | Heart Failure | 23 | Hearing Impairment/Aid |
| 8 | Hypertension | 24 | Peripheral Neuropathy |
| 9 | Kidney Disease | 25 | Vision Impairment |
| 10 | Liver Disease or Cirrhosis | **Cognition and Mood:** |  |
| 11 | Lung disease | 26 | Dementias |
| 12 | Thyroid Disease | 27 | Anxiety |
| 13 | Osteoporosis or pathological fracture | 28 | Mood Disorders (Depression, Bipolar Disorder) |
| 14 | Incontinence | **Other:** |  |
| **Function:** |  | 29 | Chronic Pain |
| 15 | Arthritis | 30 | Failure to thrive |
| 16 | Use of Durable Medical Equipment | 31 | Weight loss in the past year |

## Notes

### Competing Interest Statement

The authors have declared no competing interest.

### Funding Statement

The author(s) received no specific funding for this work.

### Author Declarations

This study was approved by the Institutional Review Board of the Miami Veterans Affairs Healthcare System (4 June 2021, reference number 1592780-1). The IRB granted a waiver for informed consent. The data was fully anonymized according to VHA standards prior to analysis.

